# Association of COVID-19-imposed lockdowns and online searches for toothache in Iran

**DOI:** 10.1101/2020.08.06.20160515

**Authors:** Ahmad Sofi-Mahmudi, Erfan Shamsoddin, Peyman Ghasemi, Ali Mehrabi Bahar, Mansour Shaban Azad, Ghasem Sadeghi

## Abstract

**Objective:** To assess the association between the lockdowns due to COVID-19 and online searches for toothache in Iran using Google Trends (GT).

**Methods:** We investigated GT online searches for the search term 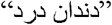 within the past five years. The time frame for data gathering was considered as the initiation and end dates of lockdown in Iran. Relative search volumes (RSVs) for online Google Search queries in 2019 was considered as the control. We performed one-way ANOVA statistical test to identify whether there is a statistical difference for RSV scores between the year 2020 and 2016-2019 for the whole country. Then we investigated the possible association of RSVs in provinces with dentists’ density, prevalence of current daily smokers, Human Development Index (HDI), Internet access, and fluoride concentration in water with linear regression. A p-value<0.05 was considered as statistically significant.

**Results:** When comparing 2020 with previous four years, there is a statistically significant difference between RSVs of 2020 with all previous years combined and each of these years (P<0.001 for all of them). In the linear model for the year 2020, HDI (B=-3.29, 95% CI: (−5.80, - 0.78), P=0.012), fluoride concentration (B=-0.13, 95% CI: (−0.24, -0.03), P=0.017), and prevalence of daily smokers (B=0.33, 95% CI: (0.13, 0.53), P=0.002) were significantly associated with RSVs. These covariates were not statistically significant for other years, except for Internet access in 2016 (B=-1.13, 95% CI: (−2.26, 0.00), P=0.050).

**Conclusion:** The RSVs for toothache in 2020 have significantly increased due to COVID-19-imposed lockdowns compared to the same period of the year in the past four years. Knowing that this period mostly overlaps with the national holidays of Nowruz in Iran, reinforces the impacts of lockdowns on people’s CSB about the toothache. In the subnational scale, the RSVs were significantly correlated with HDI, fluoride concentration, and prevalence of daily smokers which emphasises the role of socioeconomic factors in dental health and care-seeking behaviour.

## Introduction

While the outbreak of novel coronavirus disease-2019 (COVID-19) has afflicted the populations in many aspects globally, noticing its implicit consequences is of utmost importance. This can include socioeconomic, political, cultural, or even clinical aspects which could harm equitable access to healthcare during public health emergencies (PHE).^1^ Providing the best possible therapeutic clinical care could be mentioned as a critical duty for the healthcare systems in each country or region in the era of COVID-19.^2,14^ Setting urgent medical centres to provide necessary medical services for patients during the outbreak is a practised experience for many countries.^3,5,6^ Given that more hassles (e.g. patient overload, shortage of personal protective equipment, etc.) can rise during a PHE, the need for urgent healthcare can be of a more vivid nature.^7-9^ The extent to which each country and/or region faces these challenges is directly related to many factors, namely previous public health status, healthcare system stewardship, economic resilience of the country (labour markets’ liquidity, etc.), cultural norms, etc.^1,10-12^

The Internet can be introduced as one of the most popular sources of gaining information for patients all over the world. Meanwhile, the accessibility of online resources (media, social networks, scientific websites) is constantly increasing, and the way the general population seeks demanded information is parallelly changing. Care seeking behaviour (CSB) has been of researchers’ interest in the last years and especially during the COVID-19 pandemic.^13,14^

According to previous literature, some studies have suggested a rise in online searches for dental healthcare. Knowing that oral healthcare is significantly important due to its critical impacts on the daily life of every individual,^15,16^ the increasing trend of relative search volumes (RSVs) in popular online search engines like Google seems to be quite expectable. The way people seek information via Google is directly related to their CSB and preferred keywords for online searches. Google Trends (GT) is proven to be a valid and reliable tool for assessing the online search trends of populations in the medical field.^17,18^

Pain is proven to be one of the most important chief complaints about dental patients globally.^19,20^ Addressing patients’ needs for dental service when they are in pain (emergency treatments) should be critically concerned during the COVID-19 outbreak. Carter et al. have reported the acute pulpitis and periapical diseases as the most common complaints of adult and pediatric patients in urgent dental care centres of Newcastle Dental Hospital during COVID-19.^6^ Another study claimed the irreversible pulpitis and dental trauma to be the most frequent diagnoses in pediatric patients in an Urgent Dental Care Centre in the North East of England and North Cumbria.^21^ Iran is no exception to this, and the trend seems to be the same due to 55.5% of Internet accessibility (in 2015).^22^ Further, poor to the fair oral health status of the general population and especially among 35 - to 44-year-old adults in Iran could reinforce the need for urgent dental services in the country.^23^ There is a lack of evidence regarding the prevalence of dental pain in the general population; however, there exist some reports of dentists’ activity during the first 3-months period of COVID-19 spreading in Iran.^24^ Additionally, one round of national and several rounds of state-wide lockdowns have been enforced in Iran as a response to COVID-19 epidemic.^25^ This could have caused some challenges for each individual in receiving the dental healthcare they needed during the lockdowns. Accordingly, we aimed to assess the association between lockdown due to COVID-19 epidemic and searches for toothache using Google Trends in Iran.

## Materials and methods

### Google trends

We used Google Trends to investigate internet search activity during the lockdown due to COVID-19 epidemic in Iran. This web service determines the proportion of searches for a user-specified term among all searches performed on Google Search. It then provides a relative search volume (RSV), which is the query share of a particular term for a given location and period, normalised by the highest query share of that term over the time-series and presented on a scale from 0 to 100. ^26^ RSV is presented as “Interest” value in Google Trends website.

As Persian is the official language in Iran, we used “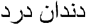” (as search term) in Google Trends, meaning “toothache” in formal Persian. There is another word for toothache in informal Persian, “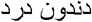”; however, since the formal term is used more widely and in many more provinces, we collected its pertaining RSVs.

Then, we gathered provincial RSVs data from the start of the lockdown in Iran (2020-03-14) to three months (12 weeks) afterwards (2020-06-14). We did the same for a similar period in 2016-2019 and chose the past year (2019) as the control event. The total number of provinces in Iran is 31.

### Statistical analysis

We plotted the trend of “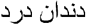” from five years ago to date. We performed one-way ANOVA statistical test to identify whether there is a statistical difference for RSV scores between the year 2020 and 2016-2019 for the whole country after normalising the data based on the Internet penetration rate. Then we investigated the possible association of RSVs in provinces with dentists’ density in 2019 (per 100,000) (retrieved from Iran’s Ministry of Health data), the prevalence of current daily smokers in 2016 (retrieved from Iran’s STEPs 2016 study ^27^, vizit.report/panel/steps/en/main.html), Human Development Index (HDI) in 2016 ^28^, Internet access in 2019 (retrieved from Iran’s Ministry of Information and Communications Technology, mis.ito.gov.ir/ictindex/), and fluoride concentration in water in 2015 ^29^ with linear regression. Statistical analyses were done using Python v3.6.7 (2018-10-20) (Python Software Foundation, Delaware, United States. http://www.python.org) on Google Colab. A p-value<0.05 was considered as statistically significant.

### Results

Overall, the RSVs trend is increasing in the past five years (Figure 1). The trend of RSVs for 2020 is increasing to a peak till the fourth week of the lockdown, then decreasing (Figure 2). When comparing 2020 with the previous four years, there is a statistically significant difference between RSVs of 2020 with all previous years combined and each of these years (P<0.001 for all of them). Figure 3 shows the RSVs for 2020 in the provinces of Iran (maps for other years are illustrated in Appendix 1.)

**Figure 1.**
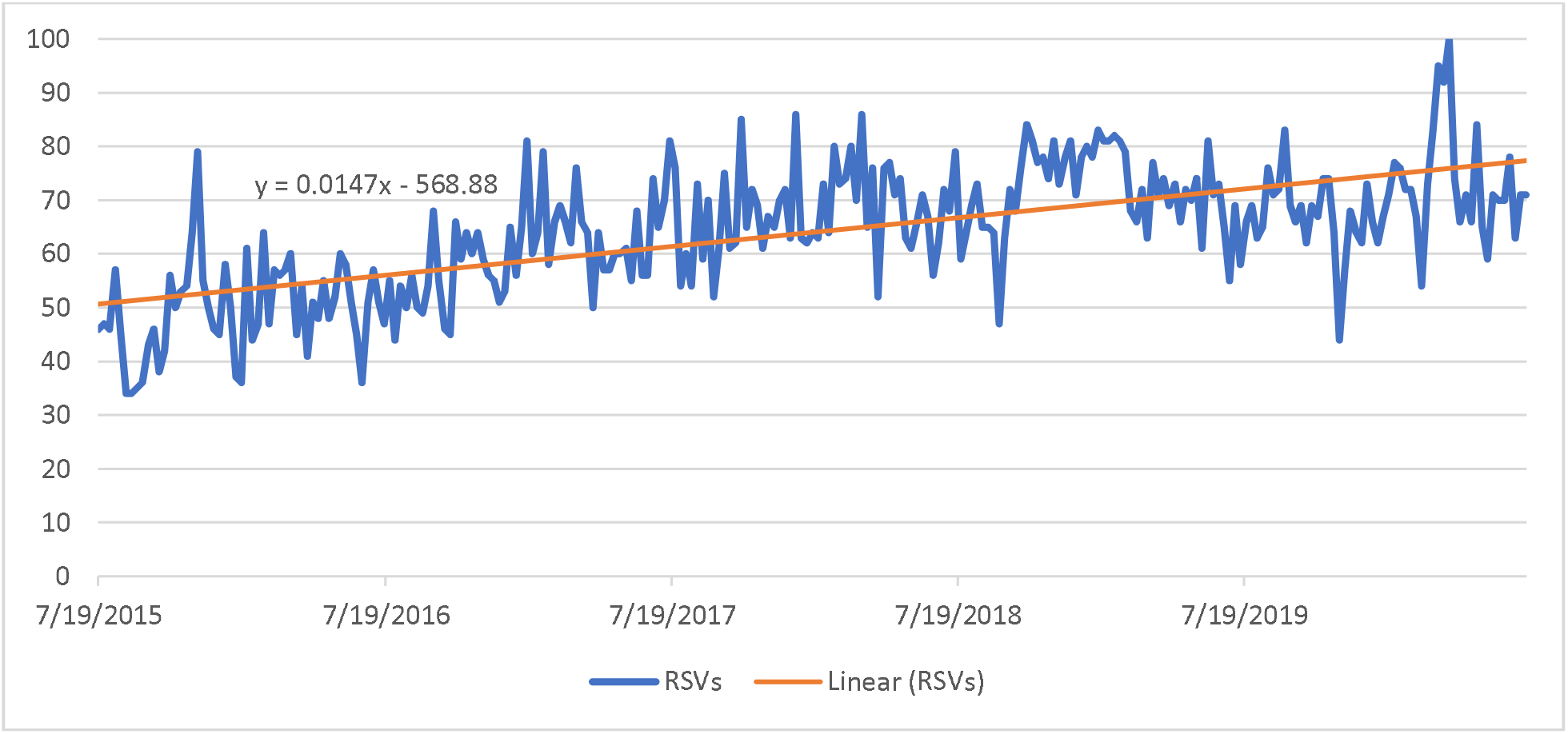
RSVs trend in past five years

**Figure 2.**
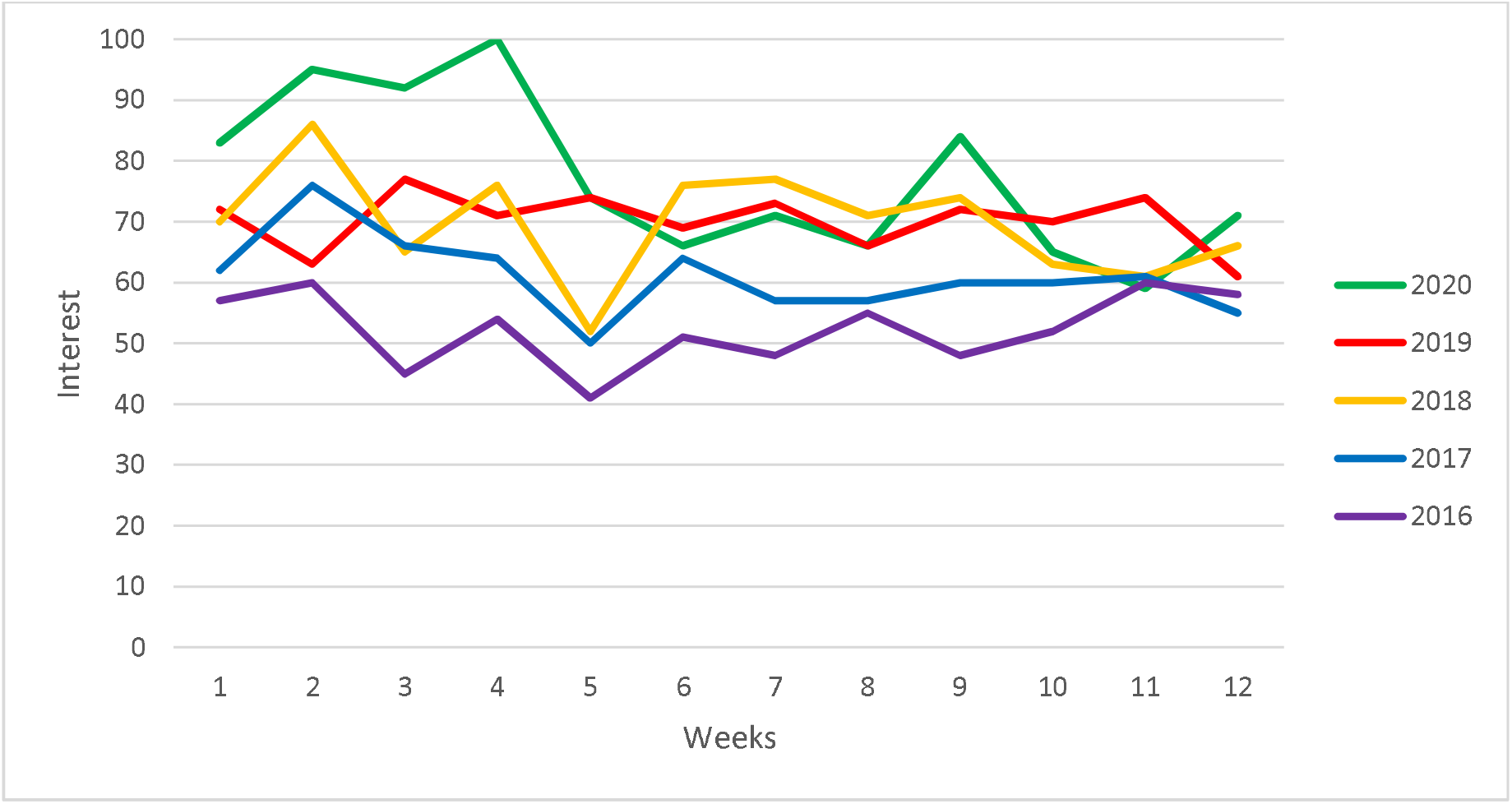
Comparing RSVs for 2020 until three months after lockdown with similar weeks in past four years

**Figure 3.**
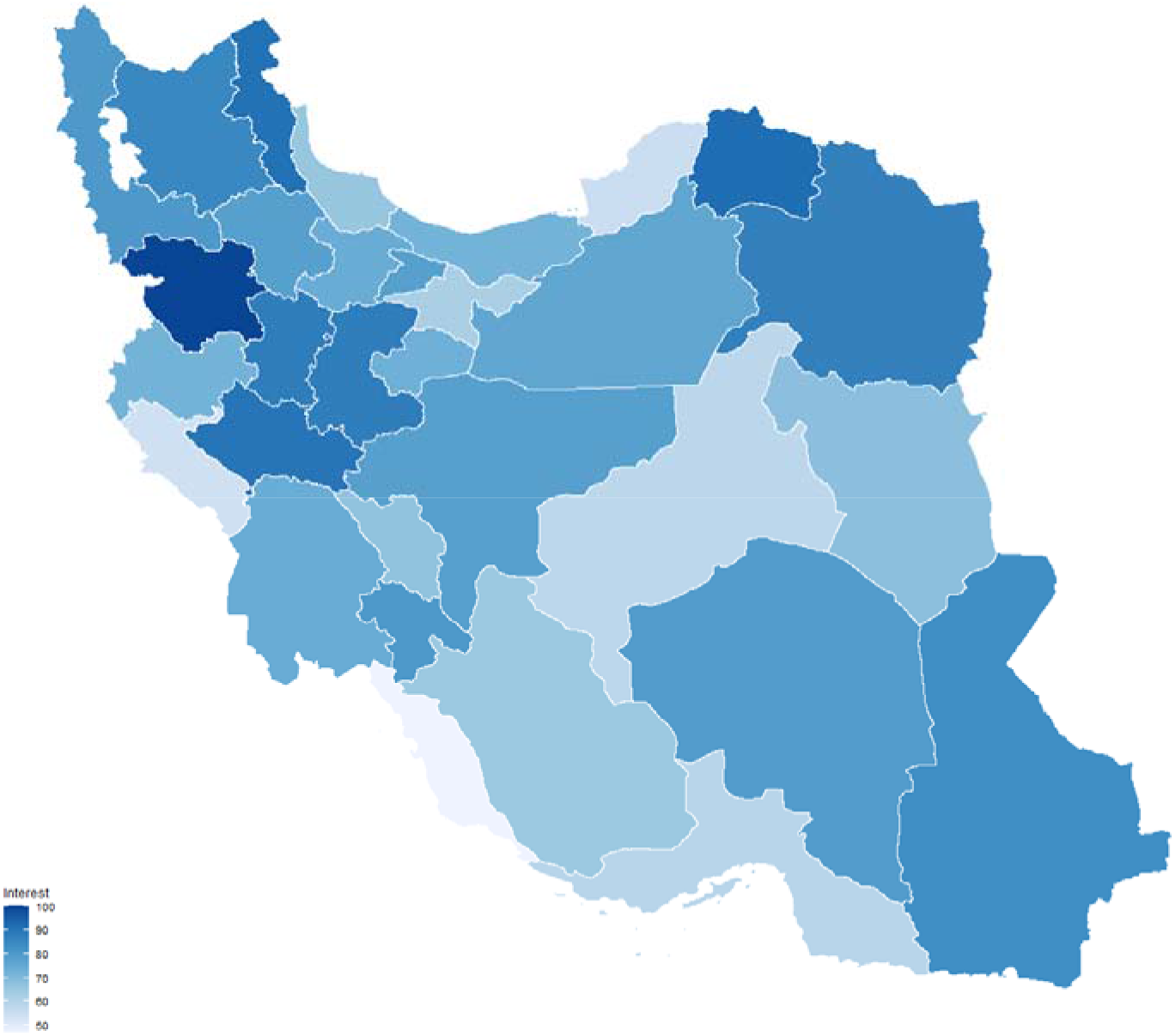
RSVs for provinces of Iran in 2020

In the linear model for the year 2020, HDI (B=-3.29, 95% CI: (−5.80, -0.78), P=0.012), fluoride concentration (B=-0.13, 95% CI: (−0.24, -0.03), P=0.017), and prevalence of daily smokers (B=0.33, 95% CI: (0.13, 0.53), P=0.002) were significantly associated with RSVs. These covariates were not statistically significant for other years, except for Internet access in 2016 (B=-1.13, 95% CI: (−2.26, 0.00), P=0.050). Full details of the linear models for each year are shown in Table 1. The concept map for the relationship between factors is illustrated in Figure 4.

**Table 1.** Linear regression results

|  | **2020** | **2019** | **2018** | **2017** | **2016** |
| --- | --- | --- | --- | --- | --- |
| **(Constant)** | **0.69\*\*\***<br>**(0.33,1.05)** | -0.41<br>(-1.05,0.23) | 0.05<br>(-0.55,0.65) | 0.33<br>(-0.29,0.95) | 0.002<br>(-0.55,0.55) |
| **Human Development Index (HDI)** | **-3.29\*\***<br>**(-5.80,-0.78 )** | 4.30<br>(-0.19,8.78) | 1.56<br>(-2.65,5.78) | -0.69<br>(-5.04,3.67) | 1.83<br>(-2.02,5.69) |
| **Internet access** | 0.36<br>(-0.38,1.09) | -0.59<br>(-1.90,0.72) | -0.50<br>(-1.73,0.74) | -0.22<br>(-1.49,1.06) | **-1.13\***<br>**(-2.26,0.00)** |
| **Fluoride concentration** | **-0.13\***<br>**(-0.24,-0.03)** | -0.12<br>(-0.31,0.07) | -0.16<br>(-0.33,0.02) | -0.09<br>(-0.27,0.09) | 0.08<br>(-0.08,0.24) |
| **Dentists' density** | -0.04<br>(-0.25,0.17) | -0.34<br>(-0.72,0.03) | -0.074<br>(-0.43,0.28) | 0.14<br>(-0.23,0.50) | -0.01<br>(-0.33,0.31) |
| **Daily smokers** | **0.33\*\***<br>**(0.13,0.53)** | -0.07<br>(-0.42,0.29) | -0.17<br>(-0.50,0.16) | 0.02<br>(-0.32,0.36) | 0.20<br>(-0.10,0.50) |
| **R<sup>2</sup>** | 0.541 | 0.274 | 0.234 | 0.092 | 0.320 |

95% confidence intervals in brackets (\* p<0.05, \*\* p<0.01, \*\*\* p<0.001)

**Figure 4.**
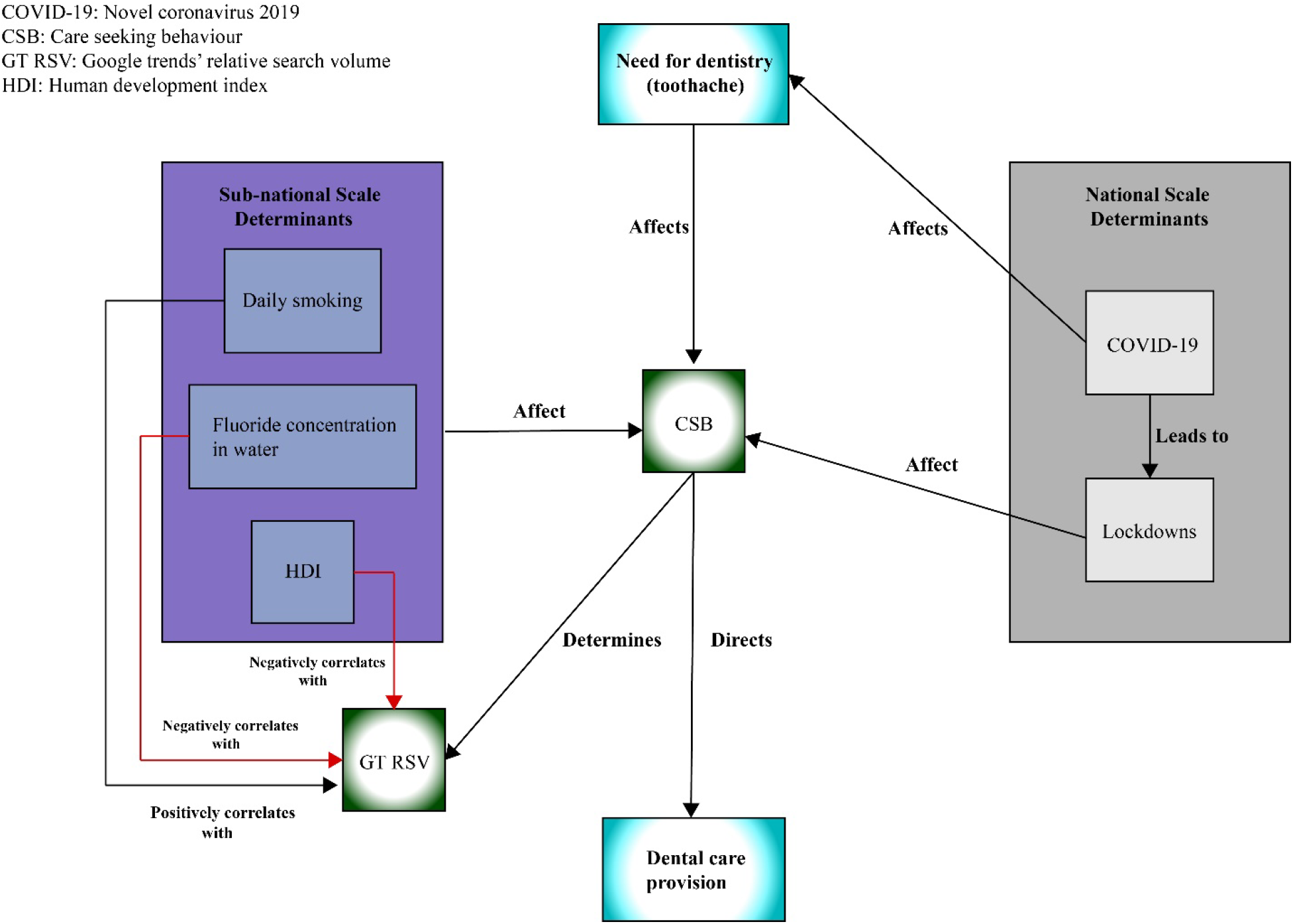
Concept map for the relationship between different factors assessed in this study

## Discussion

We found that RSVs for toothache in Iran has increased significantly during COVID-19-imposed lockdowns compared to the same period in the past four years. When analysing RSVs for provinces, we investigated whether there is a linear association between RSVs and HDI, fluoride concentration in water, and the prevalence of daily smokers. These covariates were not statistically associated with RSVs for the past four years.

While the prevalence of permanent teeth caries in Iran has reached a plateau in recent years,^15^ searching for toothache has increased. This could mean that access to dental healthcare has become more difficult. This may be true as due to worsening economic situation^30^ and significant part of dental services being provided in the private healthcare sector in Iran,^31^ people might have evicted dental healthcare from their household basket and seeking care for their toothache on the Internet. Another potential factor may be the continuously growing Persian content on the web. If we compare the Persian Wikipedia in 2016 with 2020, we will see the number of articles has increased 1.5 times.^32^ With this outburst, it seems reasonable that people get accustomed to searching for their problems before getting any help in person. This is in line with the changes that have occurred in the structure of the decision-making process from doctor’s centeredness to shared decision-making.^33^

We demonstrated a strong association between HDI and searches for toothache (B=-3.29, P=0.012). This association was not affected by the Internet penetration rate (B=0.36, P=0.325). It is now commonsense that oral diseases, including dental caries, are related to socioeconomic factors^34^, and deprived populations have worse oral health status compared to populations with better socioeconomic status. This is further in line with our results as provinces with lower HDI had higher RSVs for toothache.

Another critical factor was fluoride concentration in drinking water that provinces with higher fluoride concentrations had lower RSVs in 2020. There is a body of scientific literature that water fluoridation is a cost-effective community-based preventive approach to manage dental caries.^35,36^ However, currently, there is no water fluoridation programme in Iran, and the amount of fluoride in drinking water is different among different provinces based on the characteristics of the natural water. Southern provinces such as Bushehr have higher fluoride concentrations in drinking water^29^, and lower dental caries experiences.^37^ Fluoridation of drinking water should be considered by policy-makers to promote the oral health of the Iranians which is necessary during emergencies such as COVID-19.

It is believed that smoking is associated with several health conditions, including oral cancers,^38^ periodontal diseases,^39^ and dental caries.^40^ However, the evidence for this association with dental caries is not of high quality. This study can emphasise the effects of smoking on dental health, but we should point out that this association can be ascribed to other factors such as lifestyle habits and socio-behavioural factors.^40^

A notable finding of this study was that dentists’ density and Internet penetration rate were not significantly associated with RSVs in 2020. In recent years, the number of dental schools and dentists in Iran has been increasing dramatically.^41^ However, there are some doubts that this increase can enhance the dental health of the population. Our data cannot reject these doubts. Furthermore, it seems that Internet access is not a barrier to dental CSB in Iran.

Though the national COVID-19 lockdown mostly overlapped with the national holiday of “Nowruz” in Iran (almost lasts 15 days after 20^th^ March, every year), it seems that the lockdowns affected people’s CSB more significantly in 2020. This may be due to the closure of all dental offices during lockdowns which was unprecedented as some dental offices were open during the holidays in the previous years. Another reason might be the fear of the public for the transmission of COVID-19 in dental offices.^42^

Though, these findings should be interpreted with caution. Whilst Google shows the most market share among online search engines in Iran (87.6% as of July 2020) (gs.statcounter.com/search-engine-market-share/all/grenada), this study only assessed the CSB among Google users. Furthermore, although the Internet penetration rate is above 50% in all provinces (based on data provided in https://mis.ito.gov.ir/ictindex/provinceindex), current results may not be representative of the whole population groups. Another issue is that we could not have access to the raw data on GT; accordingly, we do not know about the times a single person has searched about toothache. This could lead to a probable error of duplication in the records (online searches).

## Conclusion

The extensive afflictions of COVID-19 shades over the field of dentistry. Care seeking behaviour of the Iranians concerning toothache, using Google search engine, shows significant differences in 2020 compared to the last four years. While investigating the considerable differences of RSVs among the provinces, we realised that HDI, fluoride concentration in the regional water and prevalence of daily smokers were all significantly correlated with the RSVs for toothache. These findings can implicitly show the importance of healthcare system performance and health policies being mirrored in general populations’ online CSB, and especially during PHEs.

## Data Availability

Data will be available upon request.

## Appendices

**Appendix 1.**
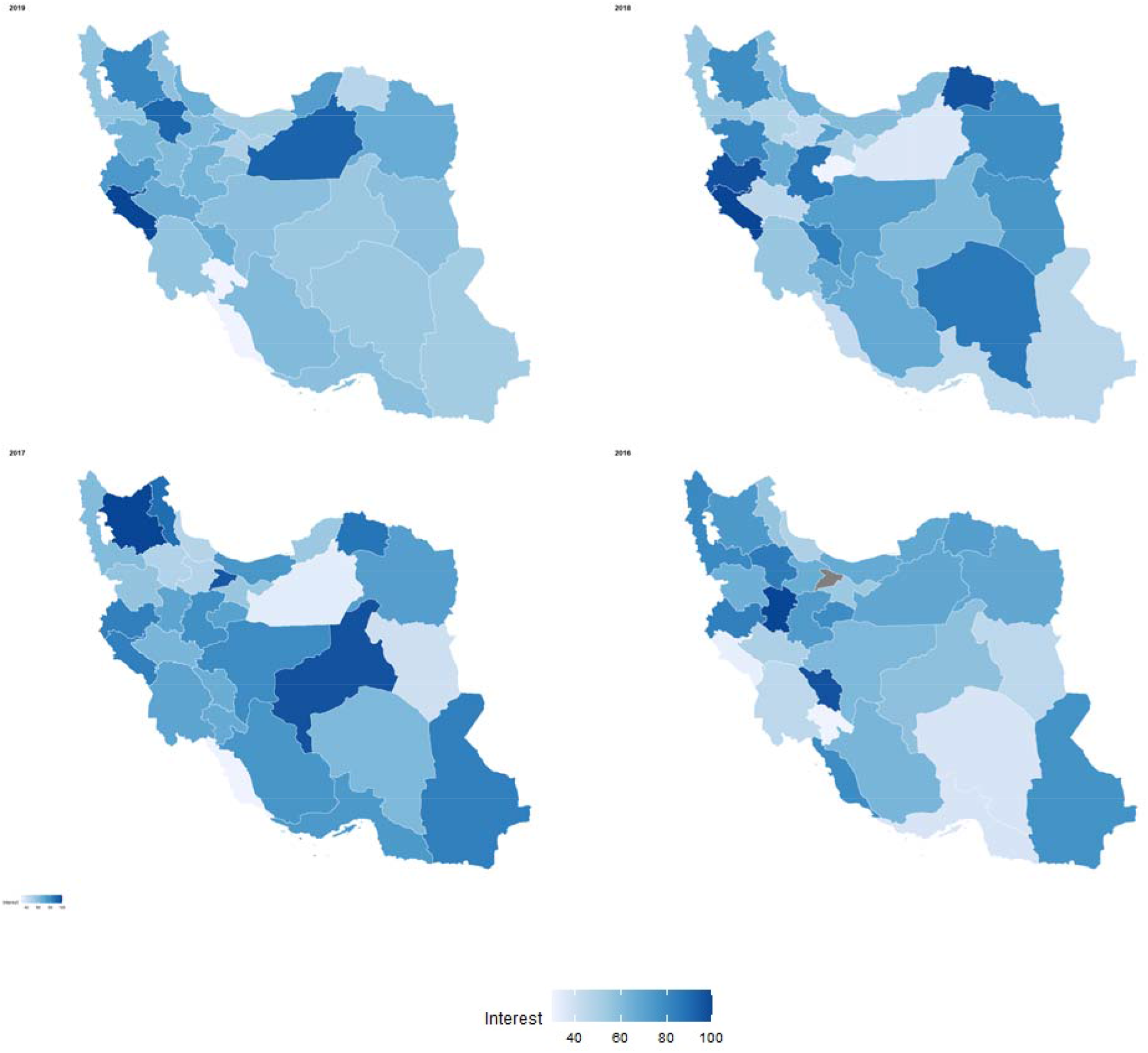
2019-2016 RSVs for provinces of Iran

## References

1. Shamsoddin E. Substantial Aspects of Health Equity During and After COVID-19 Pandemic: A Critical Review. International Network for Government Science Advice (INGSA)(2020), Policies for Equitable Access to Health (PEAH)(2020). 2020.

2. Rajan N, Joshi GP. COVID-19: Role of Ambulatory Surgery Facilities in This Global Pandemic. Anesthesia and analgesia. 2020; 131(1):31-36.

3. Provenzano DA, Sitzman BT, Florentino SA, Buterbaugh GA. Clinical and economic strategies in outpatient medical care during the COVID-19 pandemic. Regional anesthesia and pain medicine. 2020;45(8):579–585.

4. Uppal A, Silvestri DM, Siegler M, et al. Critical Care And Emergency Department Response At The Epicenter Of The COVID-19 Pandemic. Health affairs (Project Hope). 2020;39(8): 1443-1449.

5. Grasselli G, Pesenti A, Cecconi M. Critical Care Utilization for the COVID-19 Outbreak in Lombardy, Italy: Early Experience and Forecast During an Emergency Response. Jama. 2020.

6. Carter E, Currie CC, Asuni A, et al. The first six weeks - setting up a UK urgent dental care centre during the COVID-19 pandemic. British Dental Journal. 2020;228(11):842–848.

7. Patil S, Moafa IH, Bhandi S, et al. Dental care and personal protective measures for dentists and non-dental health care workers. Disease-a-month: DM. 2020:101056.

8. Mesa Vieira C, Franco OH, Gómez Restrepo C, Abel T. COVID-19: The forgotten priorities of the pandemic. Maturitas. 2020;136:38-41.

9. Lew HL, Oh-Park M, Cifu DX. The War on COVID-19 Pandemic: Role of Rehabilitation Professionals and Hospitals. American journal of physical medicine & rehabilitation. 2020;99(7):571–572.

10. Dias MC, Joyce R, Postel-Vinay F, Xu X. The challenges for labour market policy during the Covid-19 pandemic. Fiscal Studies. 2020.

11. Wong JE, Leo YS, Tan CC. COVID-19 in Singapore—current experience: critical global issues that require attention and action. Jama. 2020;323(13):1243–1244.

12. Shah J, Karimzadeh S, Al-Ahdal TMA, Mousavi SH, Zahid SU, Huy NT. COVID-19: the current situation in Afghanistan. The Lancet Global Health. 2020;8(6):e771-e772.

13. Guo H, Zhou Y, Liu X, Tan J. The impact of the COVID-19 epidemic on the utilisation of emergency dental services. Journal of Dental Sciences. 2020.

14. Hua J, Chen R, Zhao L, et al. Epidemiological features and medical care-seeking process of patients with COVID-19 in Wuhan, China. ERJ Open Research. 2020;6(2).

15. Bernabe E, Marcenes W, Hernandez CR, et al. Global, Regional, and National Levels and Trends in Burden of Oral Conditions from 1990 to 2017: A Systematic Analysis for the Global Burden of Disease 2017 Study. J Dent Res. 2020;99(4):362–373.

16. Baiju R, Peter E, Varghese N, Sivaram R. Oral health and quality of life: current concepts. Journal of clinical and diagnostic research: JCDR. 2017;11(6):ZE21.

17. Dalanon J, Matsuka Y. A 10-Year Analysis of Internet Search Trends of the Oral Health-Seeking Behaviour of Filipinos. Poverty & Public Policy. 2020;12(2):175–187.

18. Cervellin G, Comelli I, Lippi G. Is Google Trends a reliable tool for digital epidemiology? Insights from different clinical settings. J Epidemiol Glob Health. 2017;7(3):185–189.

19. Maheswaran T, Ramesh V, Krishnan A, Joseph J. Common chief complaints of patients seeking treatment in the government dental institution of Puducherry, India. J Indian Acad Dent Spec Res. 2015;2:55-58.

20. Draidi Y, AL-OLAIMAT AF, Hyasat A, Othman EF, Al Sakarna B. THE MOST COMMON CHIEF COMPLAINT AMONG JORDANIAN CHILDREN AT FIRST DENTAL VISIT. Pakistan Oral & Dental Journal. 2014;34(2).

21. Simpsons S, Sumner O, Holliday R, et al. Paediatric Dentistry and the coronavirus (COVID-19) response in the North East of England and North Cumbria. medRxiv. 2020:2020.2006.2002.20114967.

22. Iran SCo. Results of a census on accessibility and usage of information technology in Iranian households. Vol 2. Tehran, Iran: Statistical Center of Iran 2015.

23. Hessari H, Vehkalahti MM, Eghbal MJ, Murtomaa HT. Oral health among 35-to 44-year-old Iranians. Medical principles and practice: international journal of the Kuwait University Health Science Centre. 2007;16(4):280–285.

24. Sarkarat F, Tootoonchian A, Haraji A, Rastegarmoghaddam Shaldoozi H, Mostafavi M, Naghibi Sistani SMM. Evaluation of dentistry staff involvement with COVID-19 in the first 3 months of epidemiologic spreading in Iran. J Res Dent Sci. 2020;17(2):137–145.

25. Wikipedia. COVID-19 pandemic in Iran. 2020; https://en.wikipedia.org/wiki/COVID-19_pandemic_in_Iran. Accessed 8/5/2020, 2020.

26. Nuti SV, Wayda B, Ranasinghe I, et al. The use of google trends in health care research: a systematic review. Plo Sone. 2014;9(10):e109583.

27. Djalalinia S, Modirian M, Sheidaei A, et al. Protocol Design for Large-Scale Cross-Sectional Studies of Surveillance of Risk Factors of Non-Communicable Diseases in Iran: STEPs 2016. Archives of Iranian medicine. 2017;20(9):608–616.

28. Salehi MJ. Ranking Iran’s Provinces Based on Human Development and Human Capital Indices. Quarterly Journal of Research and Planning in Higher Education. 2018;24(1):27–49.

29. Taghipour N, Amini H, Mosaferi M, Yunesian M, Pourakbar M, Taghipour H. National and sub-national drinking water fluoride concentrations and prevalence of fluorosis and of decayed, missed, and filled teeth in Iran from 1990 to 2015: a systematic review. Environmental science and pollution research international. 2016;23(6):5077–5098.

30. Murphy A, Abdi Z, Harirchi I, McKee M, Ahmadnezhad E. Economic sanctions and Iran’s capacity to respond to COVID-19. The Lancet Public health. 2020;5(5):e254.

31. Jadidfard MP, Yazdani S, Khoshnevisan MH. Social insurance for dental care in Iran: a developing scheme for a developing country. Oral health and dental management. 2012;11(4): 189-198.

32. Wikipedia. [Persian Wikipedia]. 2020; https://fa.wikipedia.org/wiki/%D9%88%DB%8C%DA%A9%DB%8C%E2%80%8C%D9%BE%D8%AF%DB%8C%D8%A7%DB%8C%D9%81%D8%A7%D8%B1%D8%B3%DB%8C. Accessed 8/5/2020.

33. Cotten SR, Gupta SS. Characteristics of online and offline health information seekers and factors that discriminate between them. Social science & medicine (1982). 2004;59(9): 1795-1806.

34. Peres MA, Macpherson LM, Weyant RJ, et al. Oral diseases: a global public health challenge. Lancet. 2019;394(10194):249–260.

35. Mariño R, Zaror C. Economic evaluations in water-fluoridation: a scoping review. BMC oral health. 2020;20(1):115.

36. McDonagh MS, Whiting PF, Wilson PM, et al. Systematic review of water fluoridation. BMJ (Clinical research ed). 2000;321(7265):855–859.

37. Shoaee S, Sharifi F, Parsa PG, Sofi-Mahmudi A. Dental caries among the elderly in Iran: a meta-analysis. medRxiv. 2020.

38. Sadri G, Mahjub H. Tobacco smoking and oral cancer: a meta-analysis. Journal of research in health sciences. 2007;7(1): 18-23.

39. Zee KY. Smoking and periodontal disease. Australian dental journal. 2009;54:S44-S50.

40. Benedetti G, Campus G, Strohmenger L, Lingström P. Tobacco and dental caries: a systematic review. Acta odontologica Scandinavica. 2013;71(3-4):363-371.

41. Kazemian A. Iranian Dentistry. American College of Dentists. 2018;85(1):19–22.

42. Guo H, Zhou Y, Liu X, Tan J. The impact of the COVID-19 epidemic on the utilisation of emergency dental services. J Dent Sci. 2020.

